## Supplementary material for "A prospective diagnostic evaluation of accuracy of self-taken and healthcare worker-taken swabs for rapid COVID-19 testing": Appedix Additional graphs

**Appendix:**

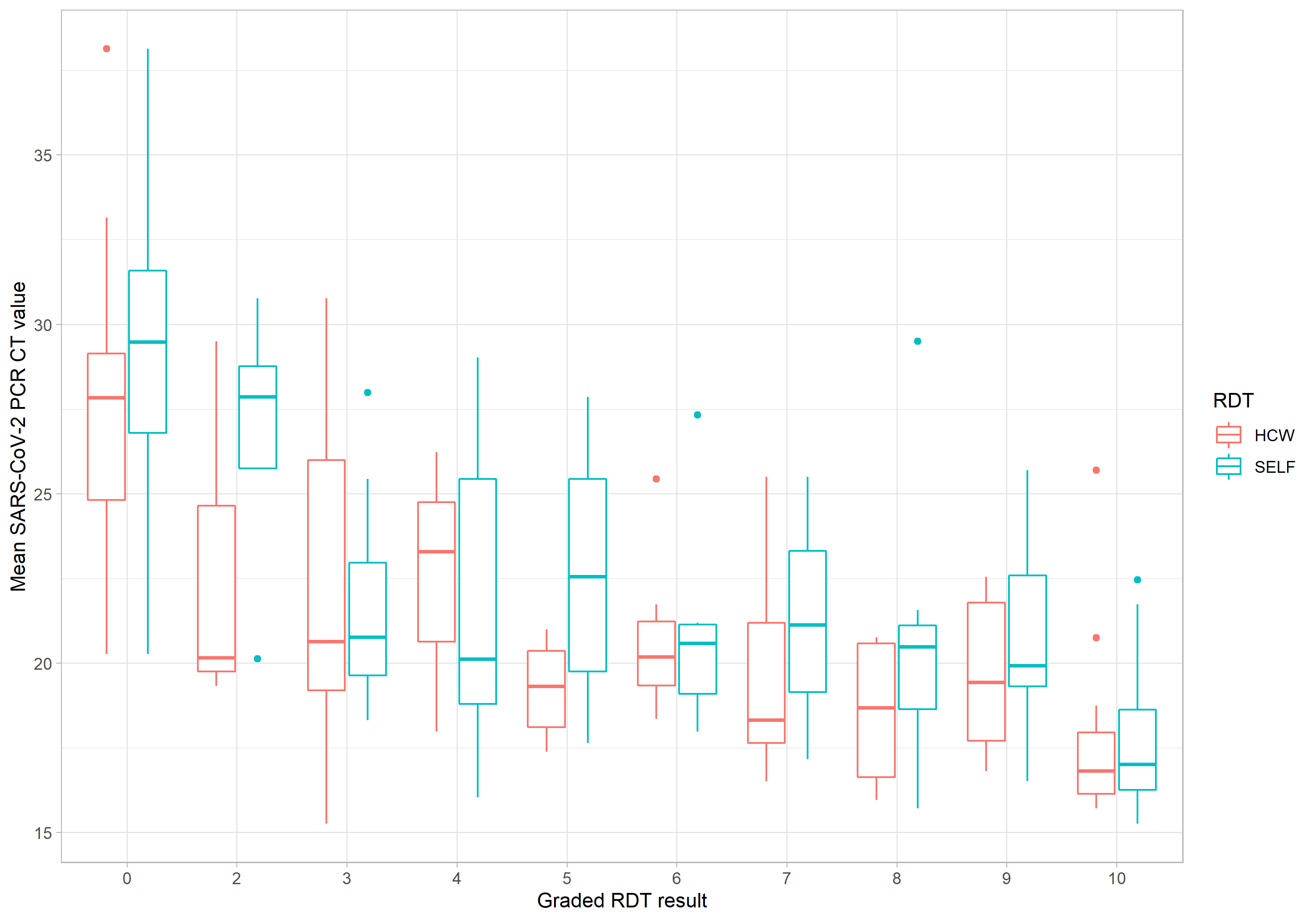
 A box plot comparing self-taken and healthcare worker taken swabs tested by Covios® RDT by mean PCR CT value.

Self-taken and HCW taken Covios® Ag RDT results by RT-PCR CT range.

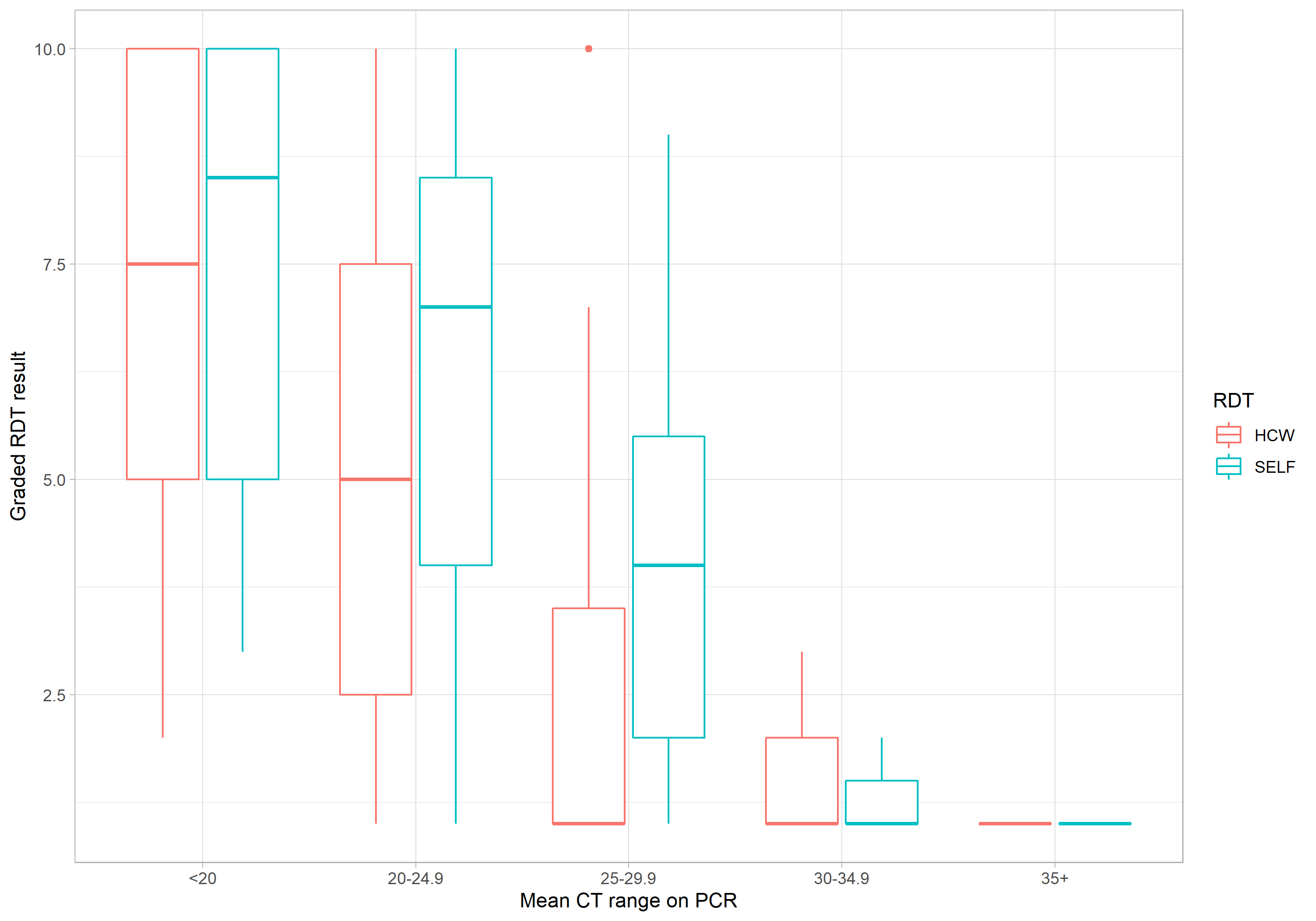
