## Supplementary figures and images for "A prospective diagnostic evaluation of accuracy of self-taken and healthcare worker-taken swabs for rapid COVID-19 testing"

### Appendix - Sampling instructions.

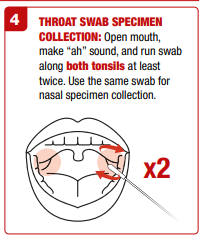

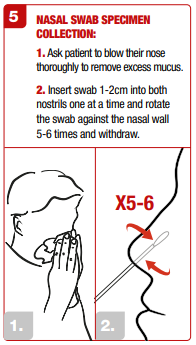

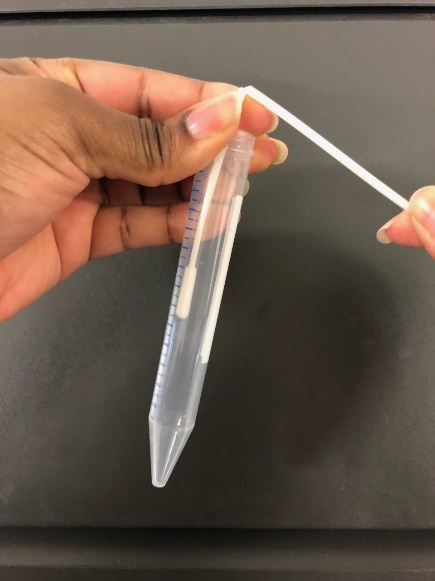

Put in green tube and snap swab in half. Screw up lid.
